# Fast Beam Scanning and Accurate Output Factor Measurements for Small-Field Dosimetry Using a Novel Scintillation Detector

**DOI:** 10.1101/2025.08.22.25334184

**Authors:** Yiding Han, Jingzhu Xu, Yao Hao, Baozhou Sun

## Abstract

**Background:** The most used instruments for small-field dosimetry have notable limitations, including the need for correction factors, limited scanning speeds, and challenges in alignment for percentage depth dose (PDD) measurements, particularly for extremely small-fields. However, plastic scintillation detectors (PSDs) are an attractive alternative for small-field dosimetry due to their correction-free nature, linear dose response, and fast response time.

**Purpose:** This study evaluates the robustness and accuracy of the dosimetric measurements using a new water-equivalent PSD in small-field dosimetry. The study also aims to report an indirect method for measuring PDD in small-fields, with a scanning time that is 5 to 10 times faster than traditional methods.

**Method:** PDDs, profiles and output factors were measured on a Varian TrueBeam 6XFFF photon beam for the field size of 0.5×0.5 cm^2^, 1×1 cm^2^, 2×2 cm^2^, 3×3 cm^2^, 4×4 cm^2^ using a new PSD from Blue Physics (BP-PSD). These measurements were compared with those obtained using a well-established PSD (Standard Imaging W2), micro-diamond (TN60019, PTW-Freiburg, Germany), and micro-silicon detectors (TN60023, PTW-Freiburg, Germany). Owing to its fast response, the BP-PSD enabled the collection of beam profiles at 31 depths, which were used to derive the PDD while avoiding detector misalignment along the beam path. Data was collected in a water tank controlled by the PTW BeamScan software. The pulse-by-pulse raw data from BP-PSD were converted to respective dosimetry data using in-house software.

**Result:** The BP-PSD demonstrated excellent agreement with other detectors for small-field output factors (FOFs), with a maximum variation of 1.6%. The BP-PSD also showed strong agreements in PDD measurements with an ion chamber (TN31013) for both 3×3 cm^2^ and 10×10 cm^2^ field sizes, achieving a 98% gamma passing rate (gamma criteria: 1mm,3%). For the profile measurements, the BP-PSD showed consistency with both the micro-diamond and micro-silicon diode detectors, with less than 1% variation in measured penumbra length. At a 3×3 cm^2^ field size, the measured penumbra length (4 mm) agreed with previously published data (3.86–4.2 mm). Additionally, for field size less than 3×3 cm^2^ the indirect PDD measurements derived from profiles showed significant improvement compared to the direct measurements using various detectors, using TPS-calculated PDD as a reference.

**Conclusion:** The BP-PSD has proven to be a robust and reliable detector for small-field dosimetry. It exhibits excellent agreement with other detectors in measuring small FOFs and provides accurate measurements with significantly faster scanning speeds in a water tank. The fast response feature enables the indirect PDD measurement method

## Introduction

In modern radiotherapy, the development of highly precise techniques such as Stereotactic Radiosurgery (SRS), Stereotactic Radiotherapy (SRT), and Stereotactic Ablative Radiotherapy (SABR) relies on the use of small photon fields to treat small volume lesions with exceptional accuracy [1]. In megavoltage photon beam dosimetry, small-fields (typically less than 4×4 cm^2^) are characterized by both the properties of the radiation beam and the specific attributes of the detectors used. From the beam’s perspective, key factors include the absence of lateral charged particle equilibrium and the partial obstruction of the primary photon source by the collimators along the beam axis. Conversely, measurement-related factors are influenced by the detector’s size and positioning relative to the beam dimensions [2–6]. The type of detectors used significantly impacts the accuracy of small-field dosimetry [4]. The International Atomic Energy Agency (IAEA) technical report series No. 483 provides guidelines for small-field dosimetry procedures and correction factors for most commercially available detectors [6].

Another challenge in small-field dosimetry is the precise alignment of the dosimeter along the beam path. Unlike large fields, small-fields lack a flat region (defined as the area with doses exceeding 80% of the central beam axis) at their center, making accurate alignment crucial, as highlighted in W. Parwaie et al. report [7]. Misalignments, such as focal spot shifts or displacements in the collimator or gantry rotation axes, can introduce substantial errors in SRS [8]. These alignment errors can significantly affect dose measurements.

Accurate dosimetry in small-fields is crucial for treatment efficacy and safety. While radiographic and radiochromic films work well, they involve lengthy processing and are orientation dependent [9–12]. Micro-diamond and micro-silicon diode detectors are widely used alternatives [6, 7].

Diamond detectors are prized for their sensitivity, tissue equivalence, and stability, with minimal impact on the radiation field. However, they are sensitive to energy and dose rate, and their performance in small-fields is debated due to issues like over-response, water equivalence concerns, under-response in fields smaller than 1×1 cm^2^, and radiation-induced charge imbalances [13–20].

Micro-silicon diode detectors are a cost-effective, highly sensitive option for relative dosimetry and beam profile measurements, but they exhibit energy, dose rate, and directional dependencies, often overestimating the dose in small-fields lacking lateral electron equilibrium due to silicon’s higher density compared to water [21–24]. Both these detectors and micro-diamond detectors, commonly used in the clinic, share several challenges in small-field dosimetry. For example, small-field output correction factors are required for TRS 483 [6]. In addition, recommends low scan speeds (resulting in long integration times per data point) for small-field sizes. Additionally, measuring percentage depth doses (PDDs) in field size < 1×1 cm^2^ is difficult due to issues with positioning accuracy and beam inclination; in extremely small-fields (<1×1 cm^2^), the detector’s vertical position may drift from the beam center, causing inaccuracies in the PDD curve at greater depths [6].

Plastic Scintillation Detectors (PSDs) have recently emerged as important tools for beam characterization and quality assurance in radiosurgery, owing to their advantageous properties such as water equivalence, high spatial resolution, energy and dose rate independence, minimal temperature response, angular independence, and linear dose response and minimal radiation damage [7, 13, 25]. The PSD detector has shown to be a correction-free detector in small-field dosimetry [6, 25, 26]. Primarily composed of doped plastic attached to an optical fiber, PSDs offer flexibility in shaping and manufacturing processes [27], enabling them to swiftly and accurately capture high-resolution dose distributions in small-fields. This capability surpasses that of traditional pinpoint ion chambers, emphasizing the critical importance of detector size in small-field dosimetry [6]. PSDs operate by converting ionizing radiation into photons within the scintillating material, which is then transmitted through optical fiber and converted into measurable electrical signals. The short fluorescence decay time of the scintillating material allows the PSDs to respond much faster than other detectors [28]. Compared to ion chambers and micro-silicon diodes, plastic scintillation detectors convert absorbed radiation into light signals within nanoseconds, highlighting their potential to significantly expedite small-field dosimetry commissioning and enable quality assurance (QA) processes.

PSDs have been extensively studied in radiotherapy applications. Early work by Beddar et al. introduced a new scintillation detector system for QA in Co-60 and high-energy therapy machines [29]. More recent studies by Carrasco et al. characterized the Exradin W1 (Standard Imaging Inc., Middleton, WI) and Jacqmin DJ et al. characterized the Exradin W2 (Standard Imaging Inc., Middleton, WI) scintillation system for radiotherapy use [30, 31]. The Blue Physics Plastic Scintillation Detector (BP-PSD, Blue Physics LLC, Lutz, FL, USA) represents another novel advancement tailored for clinical applications for small-field dosimetry. Previous investigations by Ferrer et al. [32] and Oolbekkink et al. [33] demonstrated the BP-PSD’s linear dose response, independence from dose rate, angle, and temperature, particularly on an MR-Linac. Das et al. [26] validated its consistency with other detector systems for field sizes larger than 1×1 cm^2^ for standard Linacs, but for field size = 0.5×0.5 cm^2^, significant variations were observed across detector types.

In this study, we assessed the newest generation of the Blue Physics plastic-scintillation detector, BP-PSD Model 11, leveraging its rapid-scan mode to derive percentage-depth-dose (PDD) curves in a small field (1×1 cm^2^) indirectly from central-axis profiles acquired at multiple depths in a 3-D water phantom. Detector stability was benchmarked via the output factor measurements from 0.5×0.5cm^2^ to 4×4cm^2^ with 10×10cm^2^ as reference against four reference instruments: a large-volume ionization chamber (TN31013, PTW-Freiburg, Germany), a micro-diamond detector (TN60019, PTW-Freiburg), a micro-silicon diode detector (TN60023, PTW-Freiburg), and the well-established Exradin W2 plastic-scintillation detector. To our knowledge, this is the first work to introduce and validate a rapid, profile-based technique that accurately determines PDD in small fields (1×1 cm^2^) while simultaneously minimizing errors arising from setup positioning and beam-angle (inclination) uncertainties by exploiting the BP-PSD’s fast-scan mode.

## Methods and Materials

### Blue Physics Scintillation Detector System and Readout

The BP-PSD model 11 system consists of several components: a PSD, transport optical fibers, a removable cartridge, an acquisition unit box, and Blue Physics software (BlueSoft) to visualize and analyze the data in real-time shown in [26, 34].

When a pulse of radiation interacts with the scintillation core—a combination of fluorescent dopants—it emits visible light. The intensity of light emitted by the PSD (equivalent to the number of photons produced) is proportional to the dose deposited in the detector by that pulse [35]. For BP-PSD, the cylindrical scintillation has dimensions of 1mm in diameter and 1mm in length, resulting in a 0.785mm^3^ sensitivity volume. The scintillator consists of a polystyrene core surrounded by an acrylic cladding. The core is doped with selected fluorescent compounds to optimize the scintillation response [32].

The removable cartridge houses the optical coupling for the transport optical fiber and the transducer that converts the light signal from the PSD into an electric current. It docks with the acquisition unit, which employs two interleaved integrator circuits, each with a 750-µs integration window. This dual-channel architecture guarantees gap-free sampling of pulsed beams, even when pulses straddle the end of an integration period. After each integration cycle, the acquisition unit reads and resets the accumulated charge in a capacitor. It then converts the analog signal to a digital format using an analog-to-digital converter (ADC) and sends the digital reading to the BluePhysics software (BlueSoft). The software subsequently processes these readings in real-time and generates plots.

To account for the contribution from the Cerenkov effect, BP-PSD employs a technique described by Beddar et al. [40]. The system includes a second adjacent transport fiber without a detector, identical to the fiber connected to the PSD. Essentially, BP-PSD utilizes two channels: the Sensor Channel (denoted as Rs in unit nC) captures contributions from both the PSD and Cerenkov effect, while the Cerenkov Channel (denoted as Rc in unit nC) exclusively records the Cerenkov effect. By subtracting the Cerenkov Channel from the Sensor Channel, the Cerenkov effect can be effectively removed:

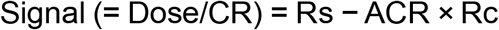

where CR is the calibration factor (cGy/nC) converting the detector signal measured in nC to cGy. This factor is determined through cross-calibration under reference conditions using a calibrated ion chamber according to established protocols.

Adjacent channel ratio (ACR) is used to correct the two adjacent channels (sensor and Cerenkov). Various methods for extracting ACR are described in [34]. In our study, we determined ACR by varying the irradiated length of the fiber while maintaining the dose at the sensor. In the first scenario, an asymmetric radiation field irradiated a very short length of the transport fibers, resulting in minimal Cerenkov light production in both the Sensor (Rs1) and Cerenkov (Rc1) channels. The actual radiation signal Signal1 is calculated as:

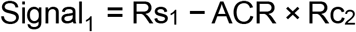

Subsequently, in the second scenario, the asymmetric radiation field was adjusted to rotate the field asymmetry and change the length of irradiated fibers while maintaining the actual dose at the sensor equivalent to the first scenario. The actual radiation signal Signal2 in this setup is:

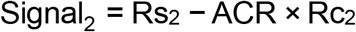

Since Signal_1_ = Signal_2_, we determined ACR using the formula:

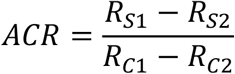

For our study, ACR was determined to be 0.963.

### Dosimetric evaluation of BP-PSD and comparison with other small-field detectors

Since the BP-PSD operates on a pulse-to-pulse basis, its raw data directly reflects the inherent fluctuations of individual linac pulses [36]. The linac output was stabilized via the daily quality check, and the dose rate constancy was maintained during this study’s measurement. To facilitate a fair comparison with detectors such as micro-silicon diode, micro-diamond, and ion chambers, which use an integrating method, we applied a rolling smoothing technique with a 40 ms window to reduce noise in the raw data. This window corresponds to an effective spatial sampling interval of 0.4 mm along the scan axis with a scanning speed of 10 mm/s and 0.8 mm with a scanning speed of 20 mm/s, whereas all reference detectors in this study were operated at a coarser spatial resolution of 1 mm at much slower scanning speed: 2 mm/s for 2×2, 3×3 cm^2^ and 1mm/s for 0.5×0.5 cm^2^, 1×1 cm^2^.

The PDD and beam profiles of the 6XFFF photon beam were measured at a source-to-surface distance (SSD) of 100 cm within the scanning water tank. The BP-PSD moved at the vendor-recommended speed of 20 mm/s and 10 mm/s during both depth dose and profile measurements for 10×10 cm^2^, and the impact of the detector speed on the beam profile and PDD was investigated. For a detailed analysis, additional measurements were performed at a scanning speed of 10 mm/s for various field sizes— from 0.5×0.5 cm^2^ up to 3×3 cm^2^ defined by the collimator jaws—with beam profiles recorded at depths of 1.3 cm, 5 cm, and 10 cm. These measurements were compared with the micro-diamond and micro-silicon diode detectors at a much slower scanning speed to confirm the BP-PSD robustness and reliability at the fast mode. Ion chamber measurements (PTW Semiflex with a volume of 0.3 cm^3^) connected to the PTW BeamScan system served as the reference standard for field size at 3×3 and 10×10 cm^2^ for checking BP-PSD accuracy. To assess the performance of the various detectors, the PDD curves were analyzed using 1D gamma passing rate: A depth point satisfies the 3 %/1 mm (2 %/2 mm) γ-criterion if a point on the reference PDD curve can be found within ± 1 mm (± 2 mm) whose dose differs by no more than ± 3 % (± 2 %) from the measured dose. 2 %/2 mm γ-criterion is a widely used criterion for dosimetry commissioning process [37, 38]. We also utilized 3% /1 mm to provide a more stringent spatial criterion. For beam profile evaluations, the penumbra was defined as the lateral distance between the 80% and 20% isodose lines, normalized to the dose at the central axis.

The indirect PDD measurement for 1×1 cm^2^ was obtained by identifying the peak of the beam profile at multiple depths. In this study, we scanned 62 beam profiles from the water surface down to a depth of 280 mm with a resolution of 1 mm to 10 mm at various depths with 10 mm/s. The total scan time took approximately 12 minutes. We compared this indirect PDD with direct PDD measurements acquired with micro diamond, micro silicon diode, and BP≥PSD detectors. The direct datasets were corrected for beam tilt using the “Beam Inclination Correction” function built into the PTW BeamScan software. The Eclipse treatment planning system v18.1 (Varian Medical Systems, Palo Alto, CA) calculation PDD served as the reference. The 1D gamma passing rate analysis, using the previously mentioned criteria, was employed for evaluation without including the build-up region (depth > 5mm).

The Field output factors (FOFs) were measured via various detectors (BP-PSD, Exradin W2, micro-diamond and micro-silicon diode detector) at two different SSD/depth (SSD = 95cm, depth = 5cm /SSD = 90cm, depth = 10cm) for field size from 0.5×0.5 to 4×4 cm^2^ defined by the multileaf collimator (MLC) with jaw size 10×10 cm^2^. A jaw-defined 10×10 cm^2^ was utilized as a reference. For small FOF measurements, this study followed the method outlined in TRS-483. The small FOF (Ω) were determined using the ratio between dose from water (*DW*) from the small-field detector (Det) at the target small-field (M*small*), and at the reference field (10×10 cm^2^, M*re f*):

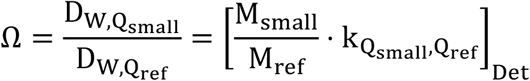

k-value 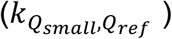, the field output correction factor, is the ratio between the beam quality correction factors of small and reference field size. For micro-diamond and micro-silicon detectors, we used the k-value based on TRS-483 [6] which collects the k-value at a small-field for Varian TrueBeam Linac. For scintillation detectors, k-value = 1 is widely accepted for PSDs [25, 26]; this factor was assumed but not independently re-verified in the present study.

It should be noted that the field output correction factor from TRS-483 doesn’t include the difference between with and without flattening filter. For the flattening filter free 6 MV beam, Underwood, et al [39] found the difference between the measured output correction factor and reported value in the TRS-483 was smaller than 1%. In the previous studies, k-value is validated via different detectors including: PSDs, micro-silicon diode, and micro-diamond detector [25, 31, 40–49].

## Results

### PDD and profile measurements

Figure 1 shows the raw and processed data for PDD and profile measurements for the 10 10 cm^2^ field size with SSD = 100 cm at 20mm/s scanning speed. It should be noted that “noisy” raw data was not from the detector but from the pulse-to-pulse fluctuation of Linac, S. Pettinato, et al. also observed via a diamond-based detector [36]. BP-PSD raw data reports the measurements for each pulse. The yellow curve represents the rolling smoothing curve with a window size of 40 ms.

**Figure 1:**
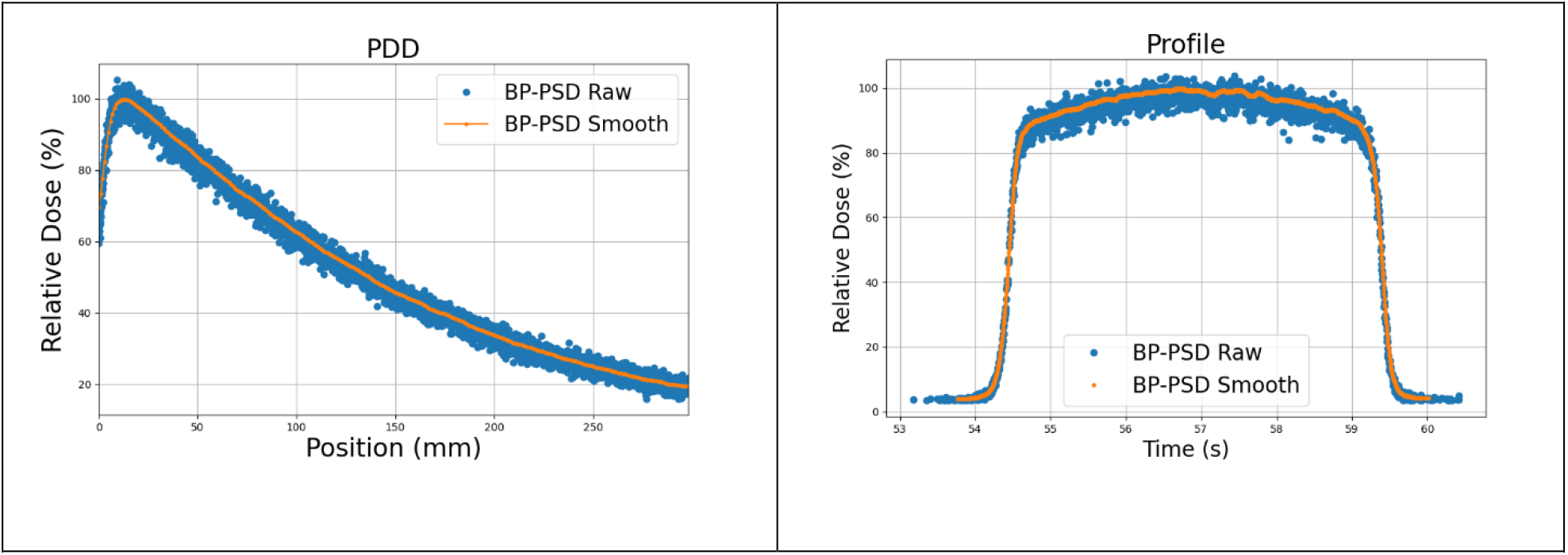
The raw and processed PDD and profile at a 10×10 cm^2^ field size. The averaging method used is a rolling smoothing with a window size of 40 ms

### Blue Physics scanning speed effect

Figure 2 shows the PDD and profile at 1.3 cm depth scanned at different velocities (20 mm/s vs. 10 mm/s) for a 3× 3 cm^2^ field size with SSD = 100 cm. The difference between the 10 mm/s and 20 mm/s scanning speeds was negligible for the PDD curve: the root-mean-square error (RMSE) between the two PDD curves is about 0.01%. However, a slight variation was observed in the penumbra region of the profile curve. The 10mm/s penumbra, 3.56 mm is slightly “sharper” than the 20mm/s penumbra, 4.40 mm. Given the minimal difference, the impact on the overall dose distribution was negligible, as demonstrated in J.A Gersh et al. study [51]. For this study, we used a scanning speed of 10 mm/s for field sizes equal to or smaller than 3× 3 cm^2^, and 20 mm/s for larger field sizes 10× 10 cm^2^.

**Figure 2:**
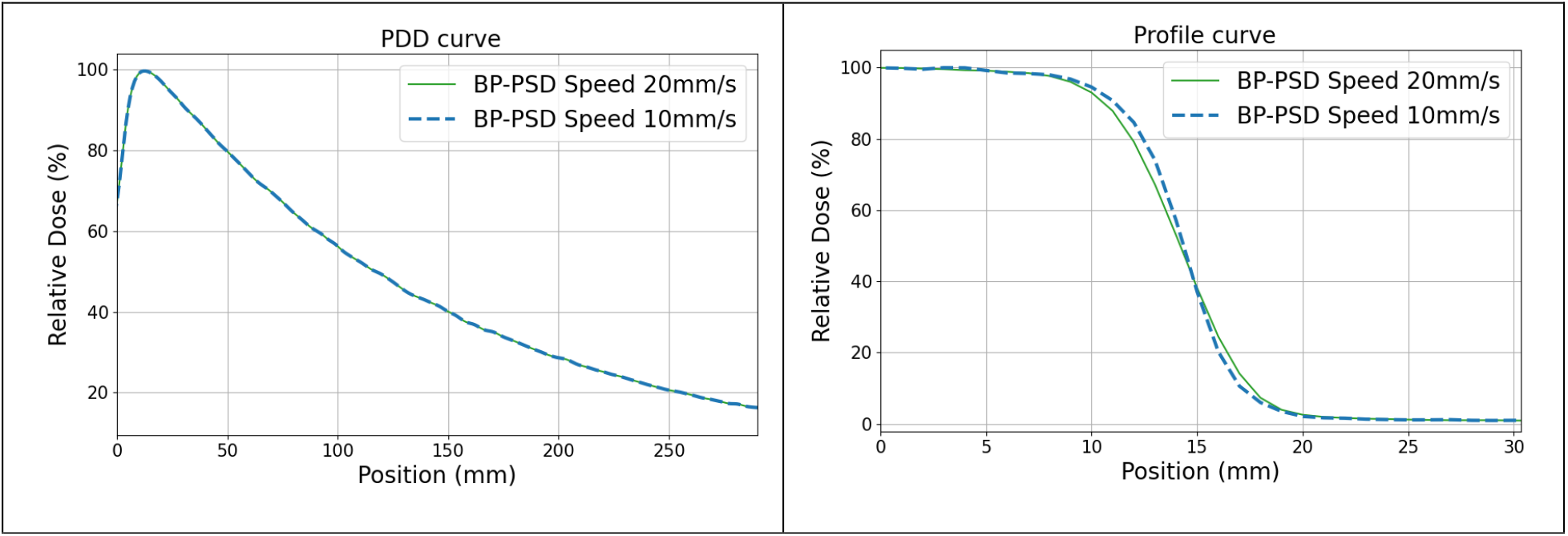
PDD (right) and profile (left) curves for field sizes of 3×3 cm^2^ at 1.3 cm depth scanned at different velocities (20 mm/s vs. 10 mm/s).

### Comparison of Percentage depth dose and beam profiles with other detectors for small-fields

Fig. 3 (a) illustrates the PDD profiles for field sizes of 3× 3 cm^2^ and 10× 10 cm^2^ with SSD = 100 cm, as measured by the BP-PSD and the ion chamber (depicted by the yellow curves, serving as the reference). The ion chamber scan was conducted at a speed of 10 mm/s, while the BP-PSD scan was performed at 20 mm/s for the 10× 10 cm^2^ field and 10 mm/s for the 3× 3 cm^2^ field, as previously described. Gamma index calculations were applied across the entire scanning depth, ranging from 0 to 298 mm, using criteria of 3% dose difference with 1 mm spatial difference and 2% dose difference with 2 mm spatial difference. The gamma passing rates were 98% for both 10 10 cm^2^ and 3× 3 cm^2^ field sizes. Fig.3(b) displays the PDD curves for a 3× 3 cm^2^ field size, measured using various detectors, including a micro-diamond detector (scanning speed of 2 mm/s), micro-silicon diode detector (scanning speed of 2 mm/s), BP-PSD (scanning speed of 10 mm/s), and an ion chamber (scanning speed of 10 mm/s). The detectors exhibit good consistency when compared to the ion chamber, which served as the ground truth, as summarized in Table 1. The position and dose criteria for the gamma indices utilized two choices: 3%/1mm and 2%/2mm. Figure 4 presents the beam profiles for a 3×3 cm^2^ field size at depths of 1.3 cm and 10 cm, measured using various detectors: ion chamber (scanning speed of 10 mm/s), BP-PSD (scanning speed of 10 mm/s), micro-diamond detector (scanning speed of 2 mm/s), and micro-silicon diode detector (scanning speed of 2 mm/s).

**Table 1:** Gamma passing rate of PDD measured by BP-PSD, micro-silicon diode and micro-diamond for a 3×3cm^2^ field size using Ion Chamber measurements as a reference.

|  | Blue Physics | micro-silicon diode | micro-diamond |
| --- | --- | --- | --- |
| Gamma passing rate<br>3%/1mm | 98% | 96% | 98% |
| Gamma passing rate<br>2%/2mm | 98% | 97% | 98% |

**Figure 3:**
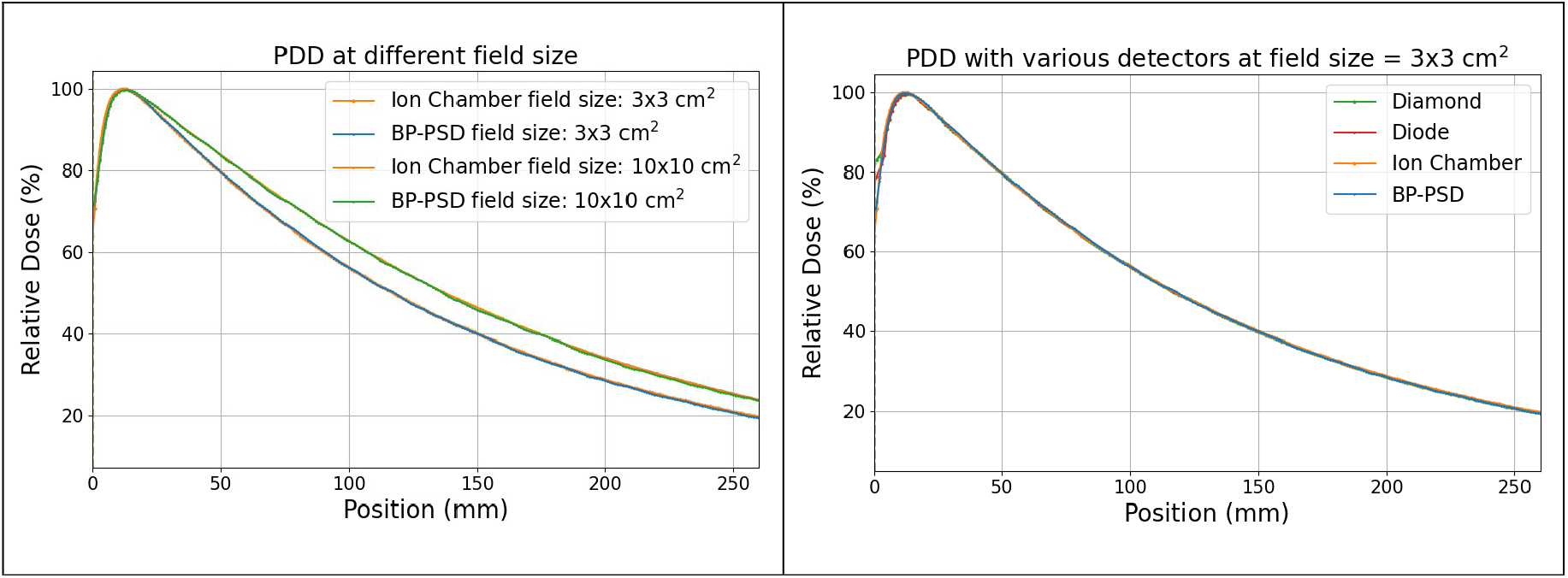
(a): PDD curves for field sizes of 10× 10 cm^2^ and 3× 3 cm^2^, showing the BP-PSD readouts post-smoothing and ion chamber measurements. (b): PDD curves for 3× 3 cm^2^ field size detected via different detectors: micro-diamond, micro-silicon diode, BP-PSD, Ion Chamber.

**Figure 4:**
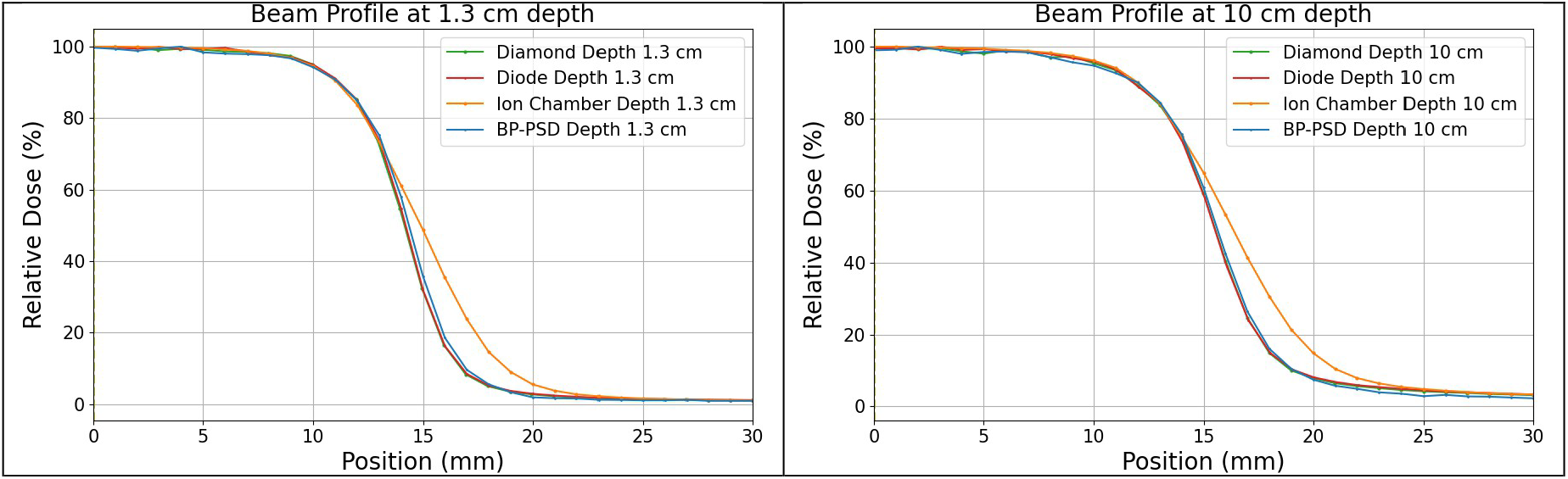
Beam profiles of 3×3 cm^2^ at different depths: 1.3 cm (a), 10 cm (b) detected via various detectors.

Figure 5 presents the beam profiles obtained from different detector measurements across various field sizes and depths, and Table 2 details the measured penumbra. The penumbra was defined as the lateral distance between the 80% and 20% isodose lines, normalized to the dose at the central axis. For BP-PSDs, the scanning speed was 10 mm/s, while for the micro-silicon diode and micro-diamond detectors, the scanning speeds were 2 mm/s for the 3 × 3 cm^2^ and 2 × 2 cm^2^ field sizes, and 1 mm/s for the 1 × 1 cm^2^ and 0.5 × 0.5 cm^2^ field sizes. The % error was defined as the deviation of the penumbra width measured with the BP-PSD from the average penumbra width obtained with the micro-silicon diode and micro-diamond detectors.

**Table 2:** Penumbra lengths (80%-20%) for different field sizes and depths measured via different detectors.

| Field Size | Depth (cm) | micro-silicon diode (mm) | micro-diamond (mm) | BP-PSD (mm) | % Error |
| --- | --- | --- | --- | --- | --- |
| 0.5x0.5 | 1.3 | 2.2 | 2.1 | 2.3 | 6.98 |
| 1x1 | 1.3 | 2.8 | 2.9 | 2.8 | 1.75 |
| 2x2 | 1.3 | 3.2 | 3.3 | 3.3 | 1.54 |
| 3x3 | 1.3 | 3.3 | 3.4 | 3.4 | 1.49 |
| 0.5x0.5 | 5.0 | 2.3 | 2.3 | 2.4 | 4.35 |
| 1x1 | 5.0 | 3.0 | 3.1 | 3.1 | 1.64 |
| 2x2 | 5.0 | 3.5 | 3.5 | 3.7 | 5.71 |
| 3x3 | 5.0 | 3.8 | 3.7 | 3.8 | 1.33 |
| 0.5x0.5 | 10.0 | 2.4 | 2.4 | 2.5 | 4.17 |
| 1x1 | 10.0 | 3.1 | 3.2 | 3.2 | 1.59 |
| 2x2 | 10.0 | 3.8 | 3.8 | 3.9 | 2.63 |
| 3x3 | 10.0 | 4.0 | 3.9 | 4.0 | 1.27 |

**Figure 5:**
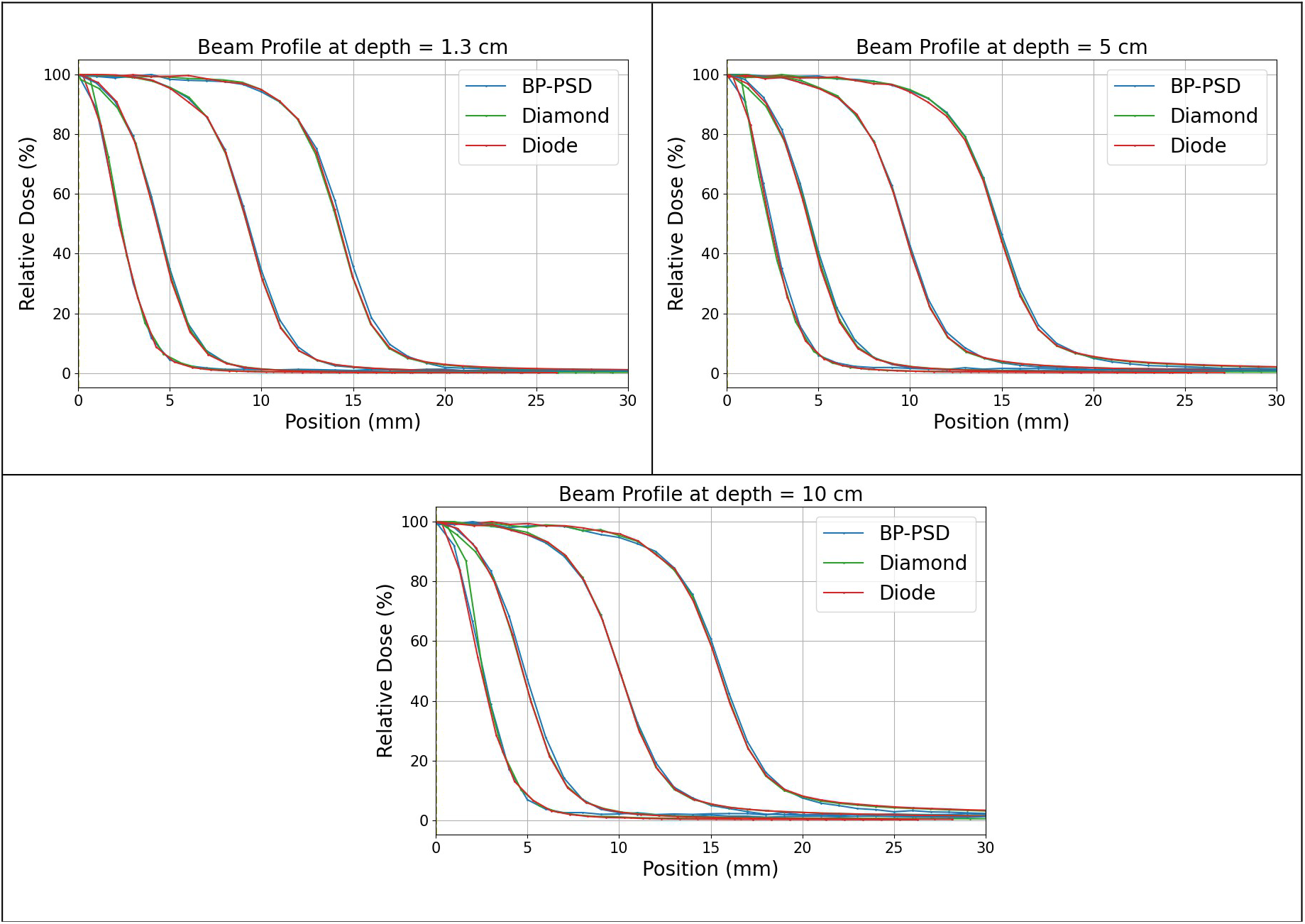
Beam profiles of 3×3, 2×2, 1×1, 0.5×0.5 cm^2^ at different depths.

In terms of time efficiency, the BP-PSD completed the following scanning tasks in less than 8 minutes: PDD, cross-plane, and in-plane beam profiles for 3×3 cm^2^, 2×2 cm^2^, and 1×1 cm^2^ field sizes, as well as beam profiles for the 0.5×0.5 cm^2^ field size. In comparison, both the micro-diamond and micro-silicon diode detectors required 40 minutes to complete the same scanning tasks.

### Indirect measurement of PDD curves

Figure 6 presents the PDD curves for the 1×1 cm^2^ field size measured using the new method, compared with measurements from different detectors and TPS-calculated PDD in a water phantom. Table 3 lists the gamma passing rates and average gamma indices for various methods excluded build-up region (depth > 5 mm), using Monte Carlo (MC) simulation with TPS data (Eclipse) results as the reference. For the gamma index calculations, the criteria were set to 1 mm (2mm) for position and 3% (2%) for dose.

**Table 3:** The gamma passing rate of different PDD measurement methods, utilizing TPS simulation as a reference.

|  | Indirect PDD using BP | Direct PDD using BP | micro-diamond PDD | micro-silicon diode PDD |
| --- | --- | --- | --- | --- |
| Gamma passing rate 3%1mm | 100% | 52% | 57% | 39% |
| Gamma passing rate 2%2mm | 100% | 43% | 47% | 34% |

**Figure 6:**
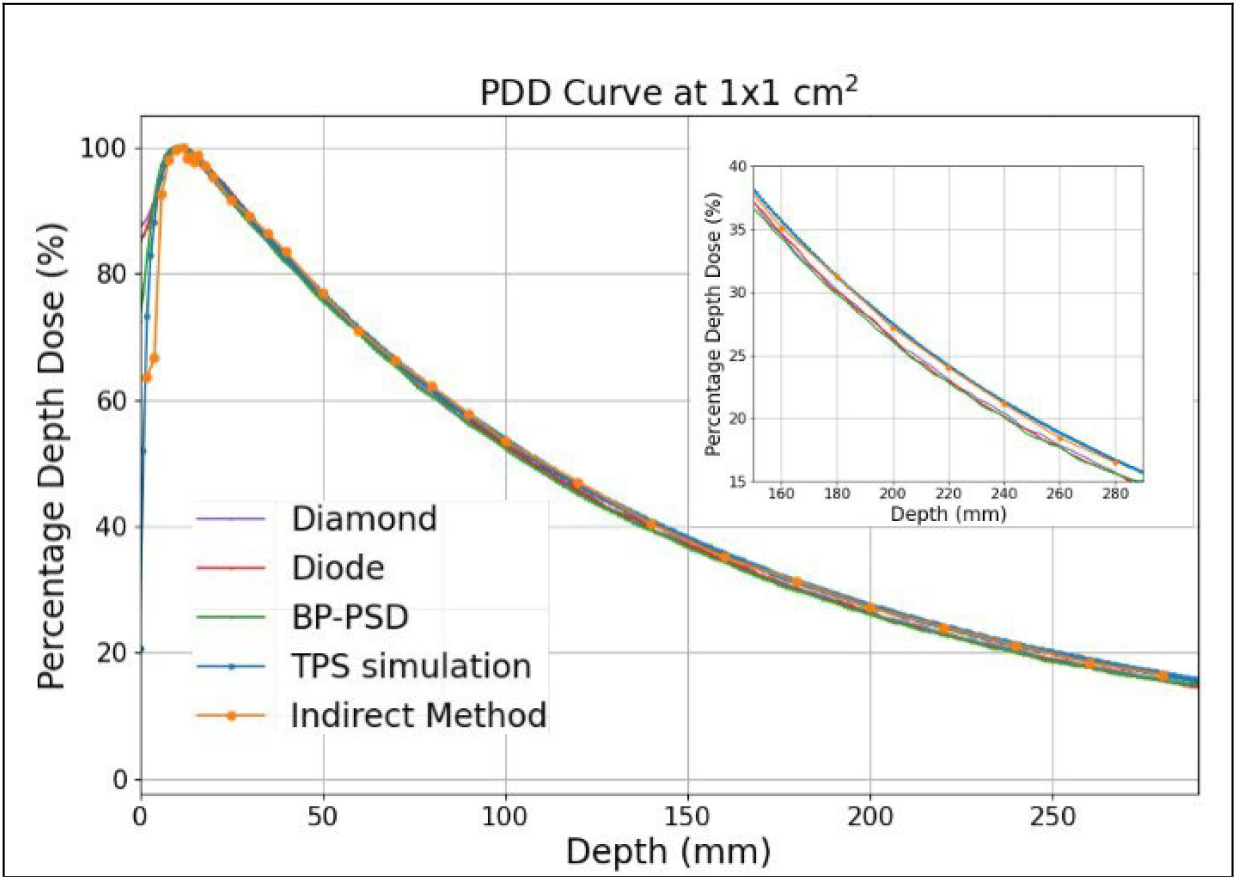
PDD curve for a 1×1 cm^2^ field size. The zoomed tail structure for showing the consistency between the TPS-calculated PDD and indirect PDD measured with BP-PSD.

### Small-field output factor

Figure 7 presents the FOF measurements at different SSD/depths (above: SSD of 95 cm with depth of 5 cm, below: SSD of 90 cm and depth of 10 cm) for small square fields ranging from 0.5×0.5 cm^2^ to 4×4 cm^2^, using a 10×10 cm^2^ reference field defined by the jaw. The red points represent measurements obtained with the BP-PSD system, while the black points correspond to those measured with the W2 detector. Both BP and W2 demonstrate excellent consistency across all field sizes, with deviations of less than 1%. Furthermore, the comparison between the BP-PSD and the micro-silicon diode detector shows a closer agreement, with differences under 0.5%. For the micro-diamond detector, the discrepancy at the smallest field size (0.5×0.5 cm^2^) was 1.6%. This variation was much smaller than the previous report result (12%) in I.J. Das et al. study [26]. For larger field sizes above 1×1 cm^2^, the variation was less than 0.6%.

**Figure 7:**
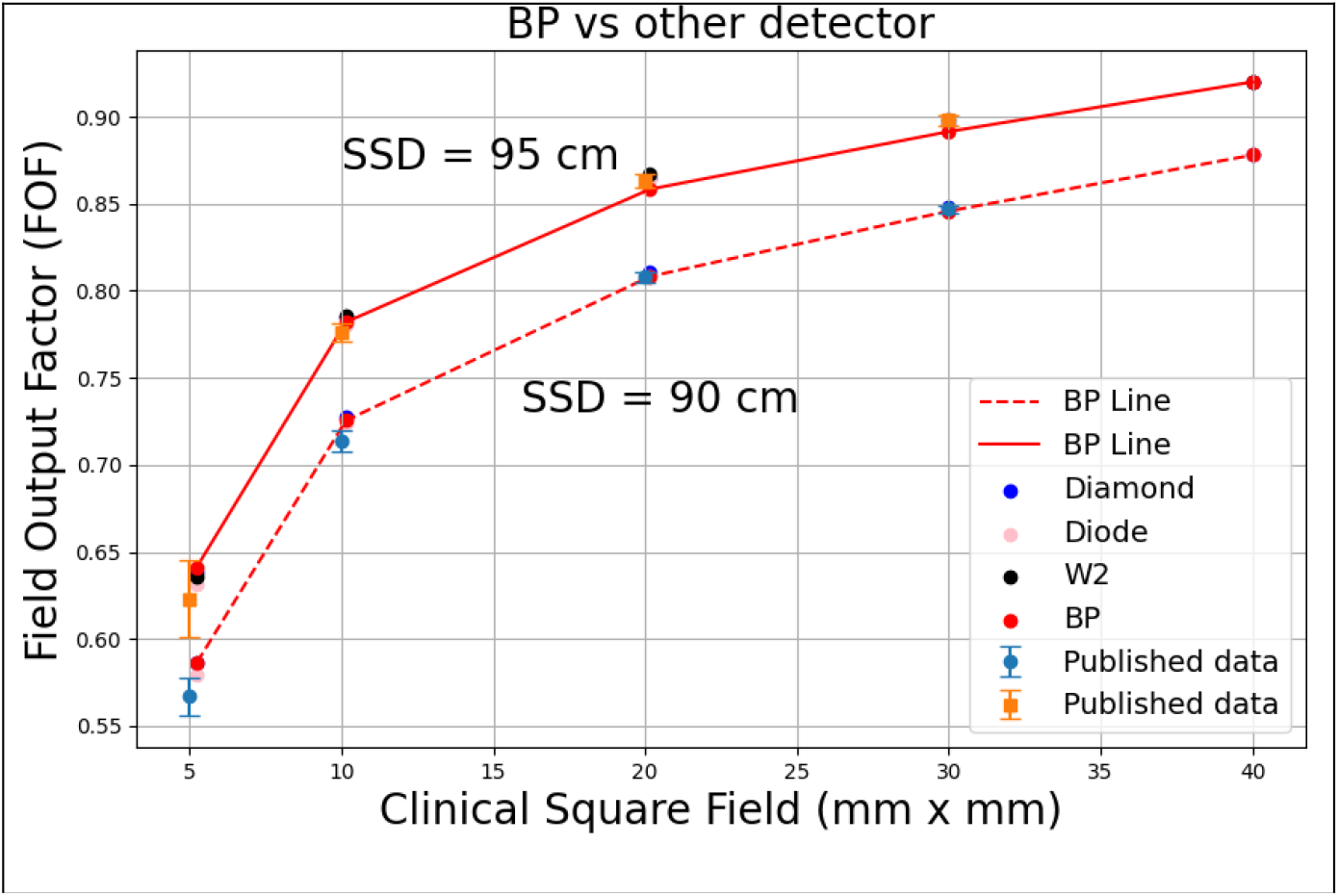
FOF in small-fields as 10×10 cm^2^ reference field at different SSDs/depths (above: SSD of 95 cm with depth of 5 cm; below: SSD of 90 cm and depth of 10 cm). Data are from various detectors: BP-PSD, Exradin W2, PTW 60019 CVD micro-diamond, PTW 60023 micro-silicon diode. The dashed line is connected through the BP-PSD points for comparison visualization.

Furthermore, these measurements were compared with the mean FOF and standard deviation for fields ranging from 0.5×0.5 cm^2^ to 3×3 cm^2^ for 6XFFF TrueBeam Varian Linac [52], as depicted in Fig. 7, the numerical comparison including standard deviation and % error is shown in Table 4.

**Table 4:**
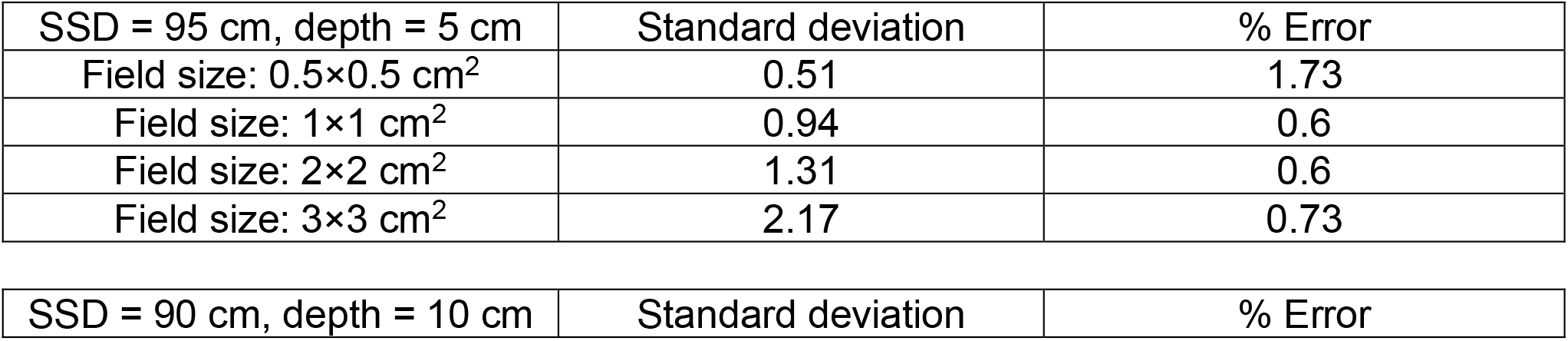
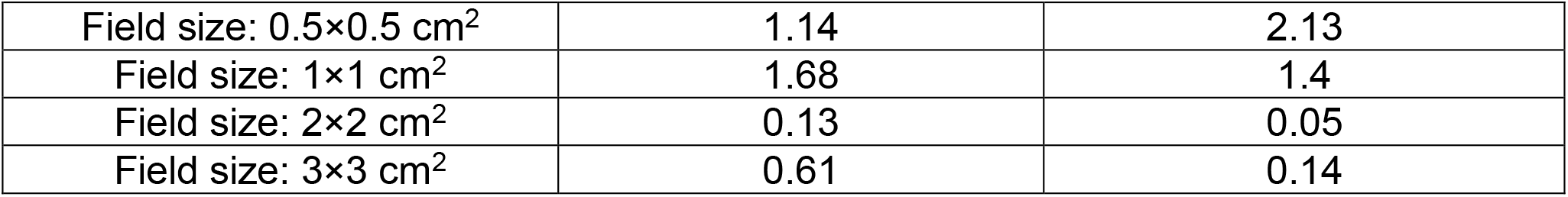
Comparison between this study BP-PSD FOF measurement with the previous literature.

## Discussion

For the PDD measurement shown in Figure 3, the observed differences via different detectors in the build-up region at a depth of 2 mm using ion chamber were as follows: BP: 0.6%, micro-silicon diode: 2.4%, and micro-diamond: 4.7%. These deviations align with the findings of a previous report [53], the micro-silicon diode showed a 4% difference, and the micro-diamond a 5% difference. For the beam profile measurement illustrated in Figure 4, the ion chamber shows a significant deviation in the penumbra region compared to the other three detectors, likely due to differences in their nominal sensitive volumes: ion chamber (0.3 cm^3^) [54], BP-PSD (0.785 mm^3^) [34], micro-diamond (0.004 mm^3^) [55], and micro-silicon diode (30 mm^3^) [56]. The impact of detector volume on measurements was well-documented in the literature, as highlighted by IJ Das, et al [57]. However, it has been reported that the penumbra length has minimal impact on clinical treatment planning systems: a penumbra length difference greater than 2 mm results in less than a 0.4% variation in dose distribution [51]. Meanwhile, the beam profile and PDD measurement via various detectors also show that fast-scanning speed with BP-PSDs achieves the consistent beam penumbras obtained from micro-silicon diode and micro-diamond detectors with a much lower speed (10 mm/s vs 1~ 2 mm/s). Consequently, a complete profile/PDD dataset across multiple field sizes can be acquired with the BP-PSD in under eight minutes, whereas the conventional detectors require roughly forty minutes.

For the penumbra of small-field size beam profile, it should be noted that both Figure 5 and Table 2 have shown a relatively large deviation, as reported from previous studies. For the 3×3 cm^2^ field size, the deviation in the measured penumbra ranges from 3.12 mm [58] to 5.76 mm [59]. The penumbra of a 3×3 cm^2^ measured using BP-PSD (4 mm) showed great consistency with both the most recent report (4.2 mm) [60] and the penumbra measured using a micro-diamond (3.86 mm) [59].

The BP-PSD’s fast-scan mode makes accurate PDD measurement feasible even for very small fields—a regime normally plagued by dosimeter positioning setup errors and beam inclination. Small fields do not feature the flat-top plateau seen in larger beams, so a slight lateral mis-positioning can shift the detector off the true central axis. Any residual beam inclination further amplifies this error because the detector may drift off axis as it moves in depth. The indirect method removes these geometric constraints. At each chosen depth, the BP-PSD sweeps laterally in a single fast pass, records the full beam profile, and pinpoints the central axis by the profile’s peak. The dose at that peak is then logged as the PDD value for that depth. Repeating this process builds a depth-dose curve that is inherently insensitive to water-tank positioning accuracy and beam inclination, enabling reliable small-field PDDs in a fraction of the usual measurement time. In this study, beam profiles were measured at multiple depths (62 profile measurements from 0 to 280 mm within 12 minutes), and the peaks of these profiles were used as points on the PDD curve. The indirect PDD method is uniquely practical with the BP PSD because its fast≥scan mode is an order of magnitude quicker than that of conventional detectors. What would require roughly two hours with a micro≥diamond or micro≥silicon diode can be completed in just 12 minutes with the BP≥PSD. As shown in Figure 6 and Table 3 this method exhibits remarkable agreement with TPS simulations in a water phantom after the build-up region, while other detector measurements show a larger deviation (< 60% gamma passing rate) even with the beam inclination geometry adjustment built within the PTW BeamScan system. Although TPS calculations are widely regarded as the gold standard for small-field dosimetry [7], the results may also have uncertainties.

For small FOF (ranging from 4×4 cm^2^ to 0.5×0.5 cm^2^), as shown in Figure 7 the BP-PSD demonstrates excellent consistency with other detectors used in this study, including the well-established PSD Exradin W2, micro-diamond, and micro-silicon diode. The variation between different detectors measured at different SSDs was smaller than 1% for field size 1×1 cm^2^. At 0.5×0.5 cm^2^, the variation is 1.6%, which was remarkably smaller than I.J Das et al. report (12%) [26]. At SSD = 95 cm, depth = 5 cm the BP-PSD agreed with published field-output-factor (FOF) data to within 0.6 % for square fields ≥ 1 × 1 cm^2^. For the 0.5 × 0.5 cm^2^ field the BP-PSD reading was 2.2 % higher than the literature mean, still well inside the reported inter-study scatter (σ = 0.51). At SSD = 90 cm, depth = 10 cm the agreement remained better than 0.6 % for fields ≥ 1 × 1 cm^2^, and the 1 × 1 cm^2^ field differed by 1.5 % (σ = 1.68). For the 0.5 × 0.5 cm^2^ field, the BP-PSD result exceeded the literature mean by 2.1 % (σ = 1.14). The larger spread for this smallest field likely stems from the absence of equivalent-square data in earlier measurements, forcing extrapolation of their reference values. These findings confirm that the BP-PSD provides accurate small-field FOF measurements; observed deviations remain consistent with the uncertainty bounds reported in previous studies.

Currently, the BP-PSD operates on a time-based data acquisition scale, whereas most scanning systems move the detector according to preset values. The beam scanning process is managed by two separate systems: the PTW BeamScan software controls the detector’s motion, while the Blue Physics software handles data acquisition. Although there is currently no direct integration between the BP-PSD and existing scanning water tank systems, such integration is under active consideration.

## Conclusion

The BP-PSD has proven to be a robust and reliable detector for small-field dosimetry. It not only shows excellent agreement with other detectors in measuring small FOF but also provides accurate measurements with significantly faster scanning speeds in a water tank. This efficiency can substantially reduce scanning time, especially when handling large volumes of tasks. Furthermore, the new BP-PSD method for PDD measurement is capable of accurately assessing extremely small-field sizes. These results demonstrate that the BP-PSD satisfies the accuracy and stability requirements for routine clinical quality assurance.

## Data Availability

All data produced in the present study are available upon reasonable request to the authors

## Notes

### Competing Interest Statement

The authors have declared no competing interest.

### Funding Statement

This study did not receive any funding

